# Assessment of Awareness and Knowledge of Human Papillomavirus Transmission and Prevention Among Tertiary Institution Students in the Plateau Central Senatorial District, Nigeria

**DOI:** 10.1101/2024.05.25.24307004

**Authors:** Juliana Rume, Imran O. Morhason-Bello, Adesina Oladokun

## Abstract

This cross-sectional study investigates awareness/knowledge about human papillomavirus (HPV) infection, transmission, prevention, and HPV vaccine among tertiary institution students in Plateau State, Nigeria. A cross-sectional study was conducted among students from two tertiary institutions in Plateau State, Nigeria. The study participants were selected using a multistage sampling technique. A well-designed questionnaire was used for data collection. Students’ responses were analysed to assess their awareness and knowledge regarding HPV transmission, prevention, and vaccination. A composite score was obtained for general HPV knowledge. A score of more than 70.0% indicated good knowledge. The distribution of the variables was examined using frequency distribution and descriptive statistics. The chi-square test was performed for bivariate analysis. Logistics regression was performed to examine the odds of having good HPV knowledge among the students. Level of significant was set at 95%.

Of 425 students in this study, 302 (71.1%) were female and 123 (28.9%) were male. There was low awareness of HPV among students, with higher awareness among the females 66 (23.1%) than the males 26 (22.2%) (p = .853). Both female 77 (26.1%) and male students 31 (26.72%) had low awareness of HPV vaccination. Among all participants, only 19 (5%) demonstrated good knowledge of HPV. Students who were employed significantly had good knowledge of HPV compared to those who were unemployed.

There is low awareness and general knowledge of HPV and its vaccination among tertiary institution students in Plateau State, Nigeria. The students’ employment status is associated with their knowledge of HPV.

## Introduction

Human papillomavirus (HPV) is a non-enveloped, double-stranded, circular DNA virus that is responsible for causing multiple epithelial lesions and cancers in both genders. It can manifest as cutaneous and anogenital warts, which depending on the subtype, may progress to carcinoma[1, 2]. HPV is predominantly transmitted through sexual contact, including vaginal, anal, and oral sex[3, 4]. Multiple health problems, such as genital warts, and cancers of the mouth/throat, cervix and anogenital sites are due to persistence of HPV infections [4]. Non-sexual skin contact and vertical transmission from mothers to babies during childbirth are other known modes of transmission[4, 5]. Generally, HPV infections are classified into high or low risk HPV depending on their oncogenic potentials. Anogenital and cutaneous warts are caused by low-risk HPVs (LR-HPVs), while anogenital (cervical, anal, vulvar, vaginal, and penile) cancers and oropharyngeal (tongue, tonsil, and throat) cancers are caused by high-risk HPVs (HR-HPVs) [6, 7]. Most sexually active men and women will be infected with HPV at some point in their life, and some may get it more than once[8].

HPV infection constitutes a major global public health concern due to its high prevalence, especially in women, where it is the main cause of cancer. Cervical HPV infection and related cancers have a varying geographic burden; they are primarily observed in low- and middle-income South American, African, and Asian countries. Previous studies indicated that Africa had the highest cervical HPV prevalence (21.1%), followed by Europe (14.2%), America (11.5%), and Asia (9.4%) among women with normal cervical cytology (NCC)[9-11].

Nigeria is the most populous country in Sub-Saharan Africa with estimated population of over 200 million people[12]. Numerous studies have shown high prevalence of HPV ranging between 16.1% to 68.8% among different population in Nigeria[13-17]. This prevalence is higher among younger women, particularly those under 30 years of age, and gradually decreases as women age in Nigeria[14, 17]. There is relatively little information on awareness/knowledge of people on HPV and associated morbidities in North Central region of Nigeria relative to regions in the Southern Nigeria. Public health programs to enhance young adults’ knowledge regarding HPV symptoms, causes, and prevention are also lacking. As a result, this study aims to explores awareness/knowledge of the transmission and prevention of HPV and HPV vaccine uptake among students in tertiary institutions in Plateau State. This study is very important for public health in Plateau State, Nigeria, because it has potential to influence policy and programs among young people in Northcentral region and their associated socio-cultural peculiarities.

## Materials and Methods

### Study design

This cross-sectional study was conducted among students in tertiary institutions in Plateau State to assess HPV awareness and knowledge on its transmission, prevention, and vaccination.

### Study setting

Data were collected from students at two different tertiary institutions in the state: Plateau State University Bokkos (PLASU) and the Federal College of Education (FCE) Pankshin. PLASU is a degree-awarding institution, while FCE is a teacher-training institution that awards both a first degree in education and a Nigerian Certificate of Education (NCE). These institutions were selected because their location is central on the Plateau, connecting the northern and southern parts of the state.

### Inclusion criteria

Students who have studied for at least one semester at each of the selected institutions, are between 18 and 30 years old, and signed the consent form to participate in the study.

### Exclusion criteria

Students from the two selected tertiary institutions who are within the required age range of 18-30 years but did not consent to participate, as well as students who are either above or below the required age range, were excluded from the study.

### Sampling techniques

A multistage sampling approach was used to select the study participants. In the first stage, PLASU and FCE were randomly selected from the list of tertiary institutions in Plateau State. Students from various faculties and departments within the selected institutions were subsequently contacted randomly.

### Sample size

The required sample size was obtained using the cross-sectional study sample size formula:

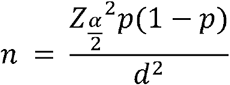

Where:

n is the sample size

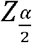 is the level of confidence (95%) = 1.96

p is the proportion of participants with awareness of HPV from the previous study = 56.4%[18] d is the precision of the study set at 5%.

The required sample size was calculated to be 379. To accommodate potential non-response biases, 12% was added to the estimated sample size, resulting in a total sample size of 425 used for the study.

### Participants recruitment and instrument for data collection

Participants were recruited through banners, student social media groups, and the students’ union government (SUG) officials. Data collection for the study was conducted at FCE between May 4th and 6th, 2023, and at PLASU between May 22nd and 24th, 2023. A self-administered questionnaire was the primary data collection tool. The questionnaires were distributed to consenting students at the institutions. The questionnaires consisted of closed-ended questions to assess the HPV awareness, knowledge of transmission, preventive practices, and vaccination status of the study students. Before data collection, the questionnaire was pretested at another higher institution in Plateau State (College of Education Gindiri) to ensure clarity and appropriateness. Necessary adjustments were made based on the results of the pilot test.

### Informed Consent

A written consent form was attached to the questionnaire which participants all read and those who are willing to participate signed prior to filling of the questionnaire. The Informed consent contained the study objectives, study procedure and assurance of confidentially and the voluntary nature of their participation. Students were also assured that data obtained will be anonymized to protect their privacy.

### Data Management and Analysis

Data collected were screened for completeness, coded, and analysed using STATA (StataCorp L.L.C.). Descriptive statistics and frequency distribution were obtained to assess the distribution of the variables. The chi-square statistical test was used to determine the association between the socio-demographic characteristics of the students and their awareness and knowledge of HPV and its vaccination. Logistics regression was performed to examine the odds of having good HPV knowledge among the students. The missing data were included in the descriptive analyses but were excluded from the tests of associations and multivariable models. For bivariate analysis, a composite score was computed. The assessment of HPV knowledge involved questions covering the mode of transmission of HPV, whether HPV can cause cervical cancer, and methods of HPV infections prevention. Each correct answer was assigned a score of 1, while an incorrect answer received a score of 0. The total score was converted to a percentage and classified as follows: a score of less than or equal to 70.0% indicated a poor level of knowledge, while a score higher than 70% indicated a good level of knowledge. A p-value less than 0.05 was considered statistically significant.

### Ethical Consideration

Ethics Committee (EC) of the Institute for Advanced Medical Research and Training (IAMRAT), College of Medicine, University College Hospital, University of Ibadan, Nigeria, gave ethical approval for this work, with reference number UI/EC/22/0028. Administrative clearance was obtained from the Plateau State Ministry of Health and the Plateau State Ministry for Higher Education. Father, permission was granted by FCE in Pankshin and PLASU in Bokkos, Plateau State, where the research data were collected.

## Results

Among the 425 students selected from the two tertiary institutions, 302 (71.1%) were female, and 123 (28.9%) were male. Students from both institutions had an average age of 23 years. Of the total students, 45 (10.6%) were married, while the majority, 380 (89.4%), were single. More than half, 289 (68.5%), of the students were unemployed, while 133 (31.5%) reported being employed. (**Table 1**).

**Table 1:** Demographic characteristics of students by institutions.

| **Variables** | **Total**<br>**n (%)** | **PLASU**<br>**n (%)** | **FCE**<br>**n (%)** |
| --- | --- | --- | --- |
| Age<sup>1</sup> [mean(sd)] | 23.3 (2.8) | 23.3 (3.1) | 23.3 (2.5) |
| **Gender** |  |  |  |
| Female | 302 (71.1) | 131 (73.2) | 171 (69.5) |
| Male | 123 (28.9) | 48 (26.8) | 75 (30.5) |
| **Educational Level<sup>2</sup>** |  |  |  |
| 100L | 110 (26.0) | 83 (46.9) | 27 (11.0) |
| 200L | 93 (22.0) | 34 (19.2) | 59 (24.0) |
| 300L | 176 (41.6) | 25 (14.1) | 151 (61.4) |
| 400L | 34 (8.0) | 34 (19.2) | 0 |
| Others | 10 (2.4) | 1 (0.6) | 9 (3.7) |
| **Marital Status** |  |  |  |
| Married | 45 (10.6) | 9 (5.0) | 36 (14.6) |
| Single | 380 (89.4) | 170 (95.0) | 210 (85.4) |
| **Religion<sup>3</sup>** |  |  |  |
| Christianity | 406 (96.7) | 174 (98.3) | 232 (95.5) |
| Islam | 14 (3.3) | 3 (1.7) | 11 (4.5) |
| **Employment<sup>4</sup>** |  |  |  |
| Yes | 133 (31.5) | 64 (36.2) | 69 (28.2) |
| No | 289 (68.5) | 113 (63.8) | 176 (71.8) |

*1 – 32 missing, 2 – 2 missing, 3 – 5 missing, 4 – 3 missing*

Generally, there was low awareness of HPV among the students, with only 92 (22.8%) indicating awareness. More female students 66 (23.1%) were aware of HPV compared to male students 26 (22.2%). University students 47 (27.2%) showed higher awareness of HPV than College of Education students 45 (19.6%). There was no discernible difference in HPV awareness between married and single students; however, employed students demonstrated more awareness of HPV than those without a job. Only 108 (26.3%) students were aware of the HPV vaccine. Both female 77 (26.10%) and male 31 (26.72%) students showed low awareness of HPV vaccination. Similar to HPV awareness, employed students 41 (31.54%) were more aware of the HPV vaccine than those unemployed 66 (23.74%). However, College of Education students 74 (31.49%) showed more awareness of the HPV vaccine than university students 34 (19.32%). The type of institution (p = 0.006) was significantly associated with awareness of HPV vaccine (**Table 2**).

**Table 2:** Awareness of HPV and its vaccination by students’ demographic characteristics.

| Variables | Awareness of HPV |  | p-value | Awareness of HPV vaccination |  | p-value |
| --- | --- | --- | --- | --- | --- | --- |
|  | Yes n (%) | No n (%) |  | Yes n (%) | No n (%) |  |
| Age [mean(sd)] | 23.8 (0.3) | 23.1 (0.2) | .063 | 23.2 (0.2) | 23.3 (0.2) | .681 |
| **Gender** |  |  | .853 |  |  | .897 |
| Female | 66 (23.1) | 220 (76.9) |  | 77 (26.1) | 218 (73.9) |  |
| Male | 26 (22.2) | 91 (77.8) |  | 31 (26.7) | 85 (73.3) |  |
| **Institution** |  |  | .072 |  |  | .006 |
| PLASU | 47 (27.2) | 126 (72.8) |  | 34 (19.3) | 142 (80.7) |  |
| FCE | 45 (19.6) | 185 (80.4) |  | 74 (31.5) | 161 (68.5) |  |
| **Educational Level** |  |  | .05 |  |  | .396 |
| 100L | 26 (24.8) | 79 (75.2) |  | 24 (21.8) | 86 (78.2) |  |
| 200L | 19 (21.1) | 71 (78.9) |  | 28 (31.8) | 60 (68.2) |  |
| 300L | 32 (19.6) | 131 (80.4) |  | 48 (28.6) | 120 (71.4) |  |
| 400L | 14 (42.4) | 19 (57.6) |  | 6 (18.3) | 27 (81.8) |  |
| Others | 1 (10.0) | 9 (90.0) |  | 2 (20.0) | 8 (80.0) |  |
| **Marital Status** |  |  | .986 |  |  | .254 |
| Married | 10 (22.7) | 34 (77.3) |  | 15 (33.3) | 30 (66.7) |  |
| Single | 82 (22.8) | 277 (77.2) |  | 93 (25.4) | 273 (74.6) |  |
| **Religion** |  |  | .221 |  |  | .615 |
| Christianity | 86 (22.2) | 301 (77.8) |  | 103 (26.1) | 291 (73.9) |  |
| Islam | 4 (36.4) | 7 (63.6) |  | 3 (25.0) | 9 (75.0) |  |
| **Employment** |  |  | .648 |  |  | .095 |
| Yes | 31 (24.4) | 96 (75.6) |  | 41 (31.5) | 89 (68.5) |  |
| No | 61 (22.3) | 212 (77.7) |  | 66 (23.7) | 212 (76.3) |  |

The students were categorized into two groups based on their composite score reflecting their general knowledge of HPV. Among all students, only 19 (5%) demonstrated good knowledge of HPV. There was no notable difference in the percentage of females (5.1%) and males (4.7%) exhibiting good knowledge of HPV. A slightly higher percentage of students from the University (6.1%) exhibited a good understanding of HPV compared to students from the College of Education (4.2%). Employed students showed a higher level of knowledge of HPV compared to unemployed students. Employment status was found to be significantly associated with students’ knowledge of HPV (p = .047) (**Table 3**).

**Figure 1:**
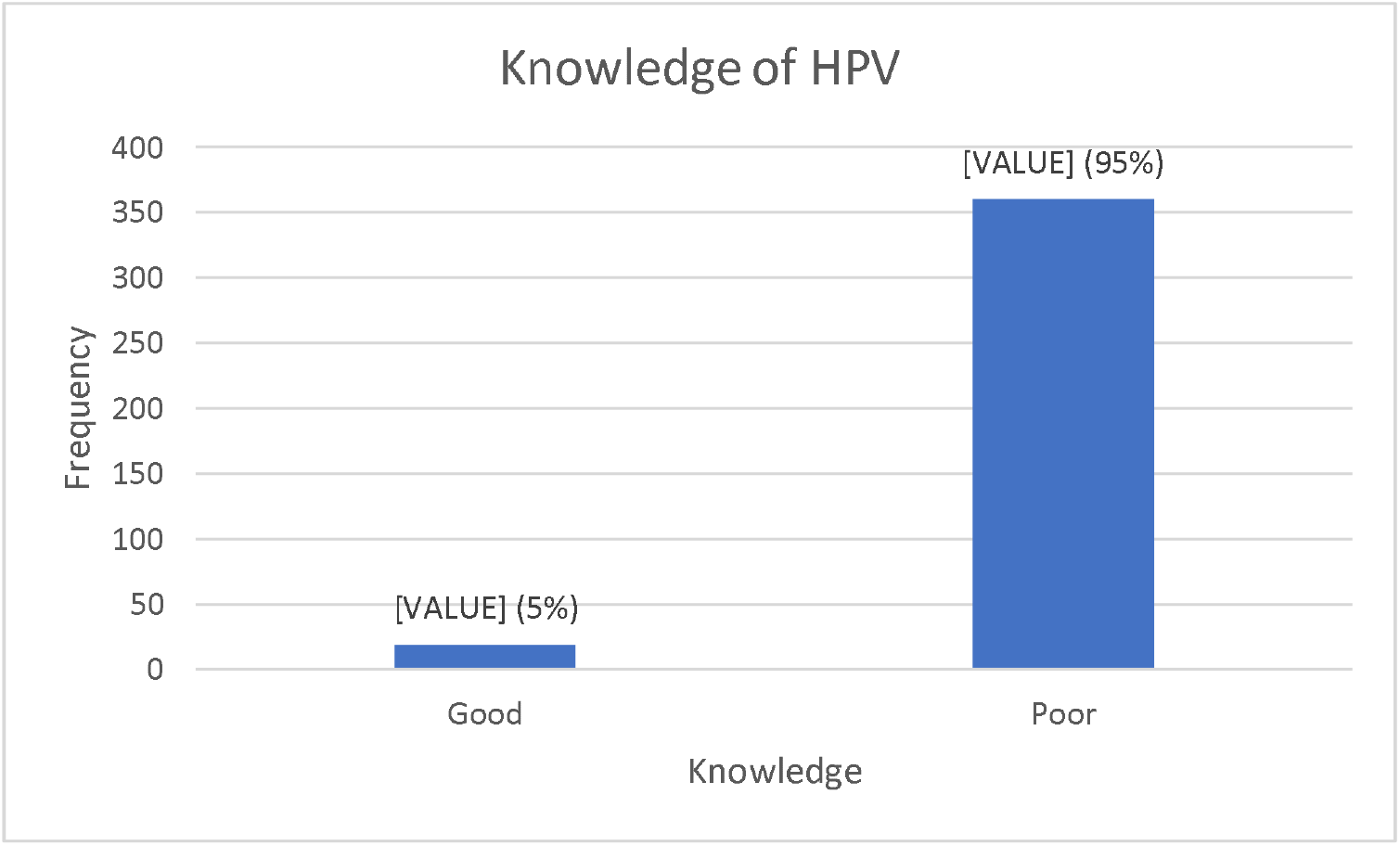
Students general knowledge of HPV

**Table 3:** General Knowledge of HPV by students’ demographic characteristics.

| Variables | General Knowledge of HPV |  | p-value |
| --- | --- | --- | --- |
|  | Poor<br>n (%) | Good<br>n (%) |  |
| Age [mean(sd)] | 23.2 (0.2) | 23.9 (0.6) | .238 |
| **Gender** |  |  | .869 |
| Female | 259 (94.9) | 14 (5.1) |  |
| Male | 101 (95.3) | 5 (4.7) |  |
| **Institution** |  |  | .412 |
| PLASU | 155 (93.9) | 10 (6.1) |  |
| FCE | 205 (95.8) | 9 (4.2) |  |
| **Educational Level** |  |  | .309 |
| 100L | 93 (92.1) | 8 (7.9) |  |
| 200L | 77 (97.5) | 2 (2.5) |  |
| 300L | 149 (96.1) | 6 (3.9) |  |
| 400L | 30 (90.9) | 3 (9.1) |  |
| Others | 9 (100) | 0 |  |
| **Marital Status** |  |  | .383 |
| Married | 39 (97.5) | 1 (2.5) |  |
| Single | 321 (94.7) | 18 (5.3) |  |
| **Religion** |  |  | .362 |
| Christianity | 348 (95.3) | 17 (4.7) |  |
| Islam | 8 (88.9) | 1 (11.1) |  |
| **Employment** |  |  | .047 |
| Yes | 111 (91.7) | 10 (8.3) |  |
| No | 246 (96.5) | 9 (3.5) |  |

The results of both crude and adjusted odds ratios of having good knowledge of HPV were presented in **Tables 4**. When adjusted for other variables, each one-year increase in age was associated with a 21% increase in the odds of having good knowledge of HPV (AOR = 1.21, 95% CI: 1.00 – 1.47). The odds of having good knowledge of HPV were lower among male students (AOR = 0.75, 95% CI: 0.22 – 2.55) compared to female students. Federal College of Education students had 1.74 times higher odds (AOR = 1.74, 95% CI: 0.49 – 6.16) of having good knowledge of HPV than University students. Furthermore, students who were employed significantly had good knowledge of HPV compared to those who were unemployed (AOR = 0.32, 95% CI: 0.11 – 0.88).

**Table 4:**
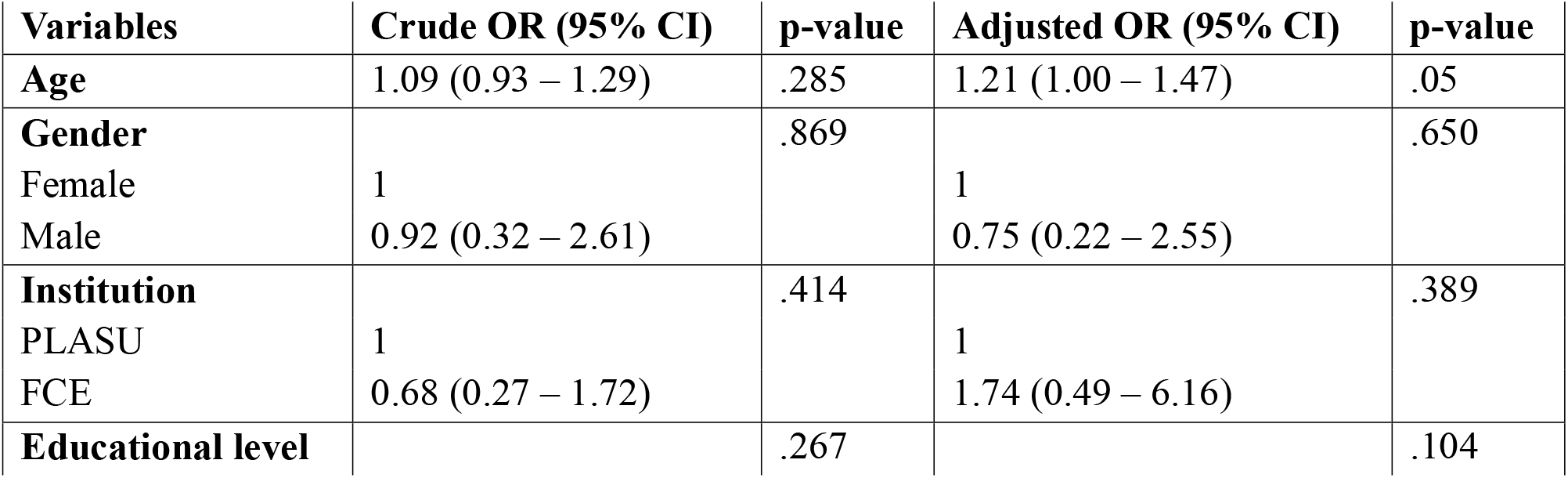
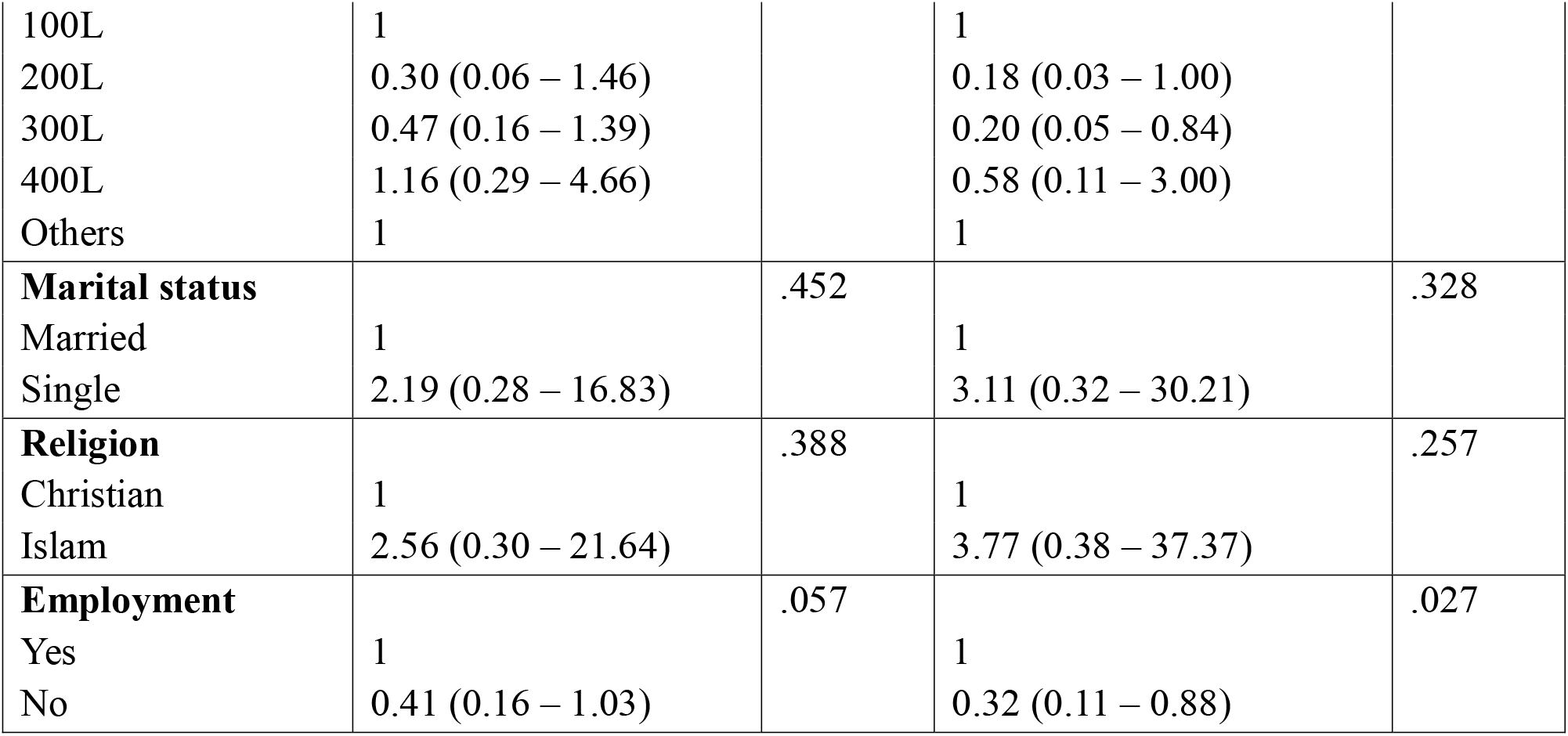
Logistic regression analysis of factors associated with good HPV knowledge.

## Discussion

This study found that there is low awareness of HPV and its vaccination among tertiary institution students in Plateau State, Nigeria. Students’ knowledge of HPV was considerably low in the study. Approximately 5% of students in tertiary institutions had good knowledge of HPV; 22.8% had ever heard of HPV; 13.9% were aware that HPV causes cervical cancer; and 26.3% knew HPV has a vaccine. The students’ employment status was found to be associated with good knowledge of HPV.

The low awareness/knowledge of HPV in this study are similar to the findings from a study by Makwe et al. conducted in Lagos and a study in Kano, where it was found that only 17.7% and 3.7% of female participants, respectively, had good awareness and knowledge of HPV [18, 19]. Given that every student had finished secondary school education, the similarity of our study’s findings to those of Makwe et al. was unexpected. The lack of state-sponsored health education initiatives about HPV and vaccination may be the cause of the low level of awareness of HPV, which is believed to be a problem in the majority of developing countries. However, further analysis revealed a significant association between educational level and HPV awareness. This association aligns with findings from studies conducted in developed countries, which also reported a relationship between increased levels of education and HPV awareness[20-22].

The prevention of HPV infections is essential for the control of HPV-related cancers. The students had extremely low levels of awareness of the HPV vaccine. Only 8.01% of students have gotten the HPV vaccine, indicating that low awareness of the vaccine and its importance has a detrimental effect on vaccine uptake. Compared to previous studies done in Benin City, Nigeria[23], there is a higher level of awareness about the HPV vaccine in Plateau state. In contrast to our study, the findings of other studies conducted in Ethiopia (31.4%)[24], Switzerland (70.7%)[25], and China (69.2%)[26] among students, showed more awareness about the HPV vaccine. Furthermore, the vaccine uptake is significantly lower than the HPV vaccine uptake recorded in developed countries like Germany (67.0%)[22]. The observed variation can be explained by differences in geographical location, government policies, healthcare activities, awareness campaigns, and socioeconomic factors. Lack of accessibility to the vaccine is another issue lowering HPV vaccination rates.

Although both genders had low scores on general knowledge of HPV, the scores were relatively lower among male students than female students. This suggests that tertiary institution students, particularly male students, may not be fully aware of HPV. This can be explained by the fact that HPV infection is primarily associated with cervical cancer in females, and the HPV vaccine is not available for males in Nigeria. Enhancing the knowledge of male students regarding HPV infection, related illnesses, and the advantages of vaccination is important.

Assessing the general knowledge of HPV among the students, we observed significant differences in the perceptions of HPV transmission and preventive techniques, particularly between genders. Most students agreed on certain modes of transmission such as unprotected intercourse and blood transfusions, but discrepancies were found in other methods. For instance, only 48% of female students and 60% of male students acknowledged French kissing as a potential mode of transmission. Furthermore, regarding preventive techniques, the study found a notable difference between both genders. For example, compared to males (76.8%), a higher percentage of female students (84.3%) considered protection during sex to be an effective preventive approach. Conversely, the majority of students, regardless of gender, recognized the importance of remaining faithful in a relationship as a preventive measure. The cultural norms surrounding health talks and specialized health efforts that focus on women’s health issues may have influenced female students, who demonstrated a remarkable knowledge of HPV preventive techniques.

This study has the following limitations: Self-report bias and recall bias may have been present in the study because we relied on self-reported data that was gathered through self-administered questionnaires. Second, because the study was cross-sectional, it is not possible to establish strong causal relationships; hence, care should be taken when interpreting the influences.

In conclusion, this study highlights the inadequate awareness and knowledge of HPV, its transmission, and preventive measures among students in Plateau State tertiary institutions. Targeted educational programs focusing on diverse educational levels and institution types are recommended to enhance HPV knowledge and promote vaccine uptake. Addressing these gaps is crucial to reducing HPV-related health risks and advancing public health efforts in the state.

## Data Availability

All data produced are available online at the link provided below.

https://kaggle.com/datasets/8fd897ac937b5a3e4bba94589b4761d828375762069358dd11371bccbfeaea2f

## Competing interest

The authors declare no competing interest in the research.

## Acknowledgment

I would like to acknowledge that the data in this manuscript are part of the corresponding author’s doctoral dissertation. I also wish to acknowledge the support of Bello Yusuf, Veronica Rume Nwankpa, Justin Clement Tangshak, and the Pan African University Life and Earth Science Institute (including Health and Agriculture) University of Ibadan.

## Authors’ contributions

Juliana Rume: Conceived and wrote the draft.

Imran O. Morhason-Bello: Revised the manuscript for important intellectual content, statistical analysis, and valuable impute.

Adesina Oladokun: Proofread and approve the manuscript for submission.

## Notes

### Competing Interest Statement

The authors have declared no competing interest.

### Funding Statement

The PhD study which this study forms a part of was supported by Pan African University, Life and Earth Science Institute (including Health and Agriculture), University of Ibadan, Oyo State, Nigeria

### Author Declarations

Ethics Committee (EC) of the Institute for Advanced Medical Research and Training (IAMRAT), College of Medicine, University College Hospital University of Ibadan, Nigeria, gave ethical approval for this work, with reference number UI/EC/22/0028.

